# Effects of medical resource capacities and intensities of public mitigation measures on outcomes of COVID-19 outbreaks

**DOI:** 10.1101/2020.04.17.20070318

**Authors:** Xia Wang, Qian Li, Xiaodan Sun, Sha He, Fan Xia, Pengfei Song, Yiming Shao, Jianhong Wu, Robert A. Cheke, Sanyi Tang, Yanni Xiao

## Abstract

The COVID-19 pandemic is complex and is developing in different ways according to the country involved. To identify the key parameters or processes that have the greatest effects on the pandemic and reveal the different progressions of epidemics in different countries, we quantified enhanced control measures and the dynamics of the production and provision of medical resources. We then nested these within a COVID-19 epidemic transmission model, which is parameterized by multi-source data. We obtained rate functions related to the intensity of mitigation measures, the effective reproduction numbers and the timings and durations of runs on medical resources, given differing control measures implemented in various countries. Increased detection rates may induce runs on medical resources and prolong their durations, depending on resource availability. Nevertheless, improving the detection rate can effectively and rapidly reduce the mortality rate, even after runs on medical resources. Combinations of multiple prevention and control strategies and timely improvement of abilities to supplement medical resources are key to effective control of the COVID-19 epidemic. A 50% reduction in comprehensive control measures would have led to the cumulative numbers of confirmed cases and deaths exceeding 590000 and 60000, respectively, by 27 March 2020 in mainland China. The proposed model can assist health authorities to predict when they will be most in need of hospital beds and equipment such as ventilators, personal protection equipment, drugs and staff.

**One sentence summary:** Multiple data sources and cross validation of a COVID-19 epidemic model, coupled with a medical resource logistic model, reveal that the key factors that affect epidemic progressions and their outbreak patterns in different countries are the type of emergency medical response to avoid runs on medical resources, especially improved detection rates, the ability to promote public health measures, and the synergistic effects of combinations of multiple prevention and control strategies.

## Main Text

In the absence of effective treatments for, or vaccines against, COVID-19, the early adoption of strict prevention and control measures will undoubtedly play a very important role in limiting the spread of the virus and the growth of the epidemic *(1-8)*. This has been shown by the successes of China, South Korea and other countries in curbing the spread of the virus and achieving important prevention and control results *(7, 8)*. For example, unprecedented restrictive measures, including travel restrictions, contact tracing, quarantine and lock-down of entire towns/cities adopted by the Chinese authorities has resulted in a significant reduction in the COVID-19 epidemic in mainland China *(7, 8)*. However, with the increase of COVID-19 cases in the world, especially the sharp increase in the cumulative number of reported deaths, the shortage of medical resources has become the most serious threat facing countries experiencing serious epidemics.

Enhancing the detection rate can lead to early quarantine or isolation of latent and/or infected individuals and then the numbers of confirmed cases *(2, 5, 8)* increase significantly. A common problem faced by many countries is to what extent can improvements to medical resources be synchronized with an increase of patient numbers, in order to avoid runs on medical resources. How to quantify the levels of improvements in prevention and control measures and of medical resources in order to reveal differences in the epidemic’s progression in different countries remain unclear, but are addressed in this study. In order to investigate this issue, we propose a general COVID-19 epidemic transmission dynamics model that includes limitations of medical resources and enhancing prevention and control strategies in six selected countries (China, South Korea, Japan, Italy, Spain and Iran) *(7, 8)*. The formulated transmission model is parameterized by multi-source data such as the numbers of newly reported cases and the cumulative numbers of deaths for each country.

Using information such as the number of beds per thousand people in each country and differences in increasing volumes of medical resources (closely related to medical staff numbers) that can be provided by each country during public health emergencies *(9-14)*, we modeled the number of beds provided by each country during the development of a COVID-19 epidemic with the logistic growth function using country-specific varying growth rates and carrying capacities (section SM1 of Supplementary material (SM)). In order to represent the limitation of hospital beds we divided confirmed cases into two groups in terms of severity of symptoms: non-hospitalized and hospitalized. The non-hospitalized individuals, who may later be admitted to hospital depending on the number of hospital beds available, can become a new source of infection, leading to new cases including family cluster infections. Hence the dynamics of hospital beds need to be nested within the transmission dynamic model to examine the level of improvement in medical resources on a COVID-19 epidemic in each country studied.

The multiple data sources including the numbers of newly reported cases and the cumulative numbers of reported deaths for all six countries were used to estimate the unknown parameters and to fit the data (Figs.1 and S2, Table S2). The parameter values associated with the intensity of disease transmission in each country are compared and discussed in SM3. Like China, the epidemic in South Korea is almost stable, and their respective effective reproduction numbers have, by mid-April 2020, both been less than 1 for six weeks (Fig.1 (B, C). The COVID-19 epidemic in Japan has been fluctuating on a small scale with a lot of random fluctuations. However, since 25 March, the epidemic has rebounded, the numbers of newly reported cases and deaths have continued to increase (Figs.1(D) and S2(C)). The Italian and Spanish epidemics seem to be about to peak and they are approaching their turning points, but their cumulative death rates will continue to rise, and there is no sign of stabilization in the short term (Figs.1(E, F) and S2(D, E)). Finally, the epidemic in Iran has a strange trend, with repeated and huge fluctuations (Figs.1(G) and S2(F)).

**Fig.1:**
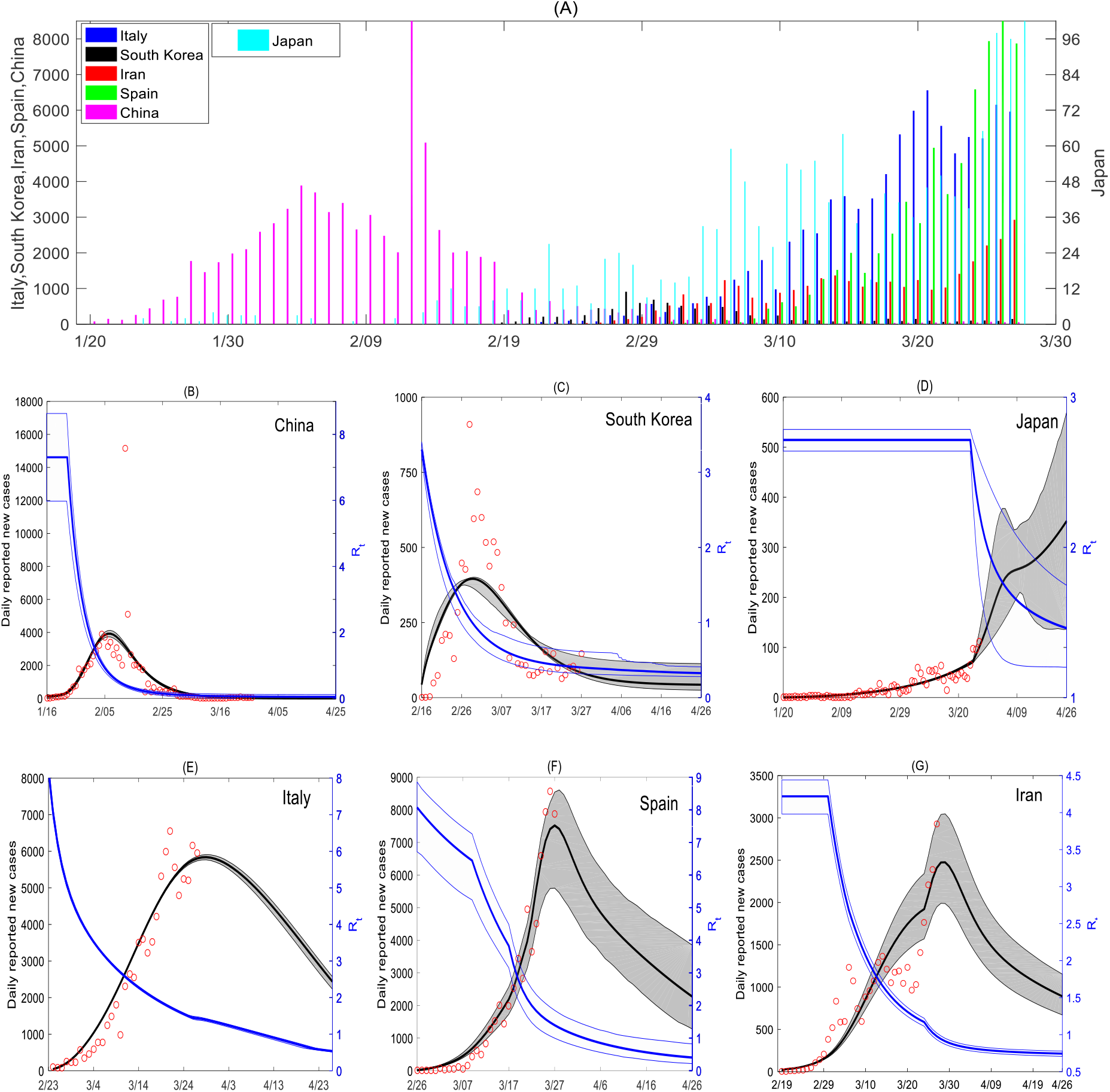
Data, curve fitting and effective reproduction numbers. (A). Numbers of newly reported cases for China, South Korea, Japan, Iran, Italy and Spain from 23 Jan to 27 March 2020. (B – G). Numbers of newly reported cases (red open circles) for China (B), South Korea (C), Japan (D), Italy (E), Spain (F) and Iran (G) and data fitting and 95% confidence intervals (gray) with predictions for one more month. Blue lines are the effective reproduction numbers *(R*_*t*_) and their 95% confidence intervals.

In order to reveal the complex patterns and huge differences in the COVID-19 epidemics among the various countries shown in Figs.1 and S2, we compared the effectiveness and timeliness of the continuously strengthened comprehensive prevention and control strategies in various countries (Fig.2), with a view to increasing our understanding and making suggestions for future prevention and control strategies. To do this, we quantified the intensities of the control measures against COVID-19 epidemics for each country by estimating the evolution of contact rate (c(t)), quarantine rate *(q(t))*, detection rates *(δ*_*I*_*(t)* and *δ*_*T*_*(t)*) and medical resource capacity *(H*_*c*_*(t)*) (see sections SM2 and SM3).

It follows from Fig.2 that five of the six countries, but not Japan, are constantly increasing their numbers of beds available *(H*_*c*_*(t)*, green curves) for COVID-19 patients with the development of the epidemic in accordance with each country’s medical capacity. The rates of increases in the numbers of beds are in the order South Korea, China, Italy, Spain and Iran. South Korea and China would soon be able to reach the maximum number of medical beds needed after their control intensity improvements (Fig.2(A) and (B)). Due to the low rate of increase of infected individuals in Japan, so far there is no urgent need to supplement the number of beds for high-risk patients there (shown in Fig.2(C)). The contact rate function (blue curve in Fig.2) in China has declined very fast since 23 January when Wuhan city and all parts of the country continued to take stringent control measures. The response speed of increasing social distancing and strengthening self-quarantine measures was very fast in mainland China, while South Korea’s, Italy’s and Spain’s contact rate functions gradually decrease. Italy and Spain have relatively low quarantine rates (red curves in Fig.2) while the other four countries all have high contact tracing followed by quarantine. Japan’s quarantine rate remains at a high level so far. The relatively high detection rates, followed by strict quarantine, are associated with quick control of the epidemic (as illustrated for China, South Korea and Iran in Fig.2 (A, B, F)) while the epidemics in Italy and Spain with low detection rates became worse (Fig.2 (D, E)).

**Fig.2:**
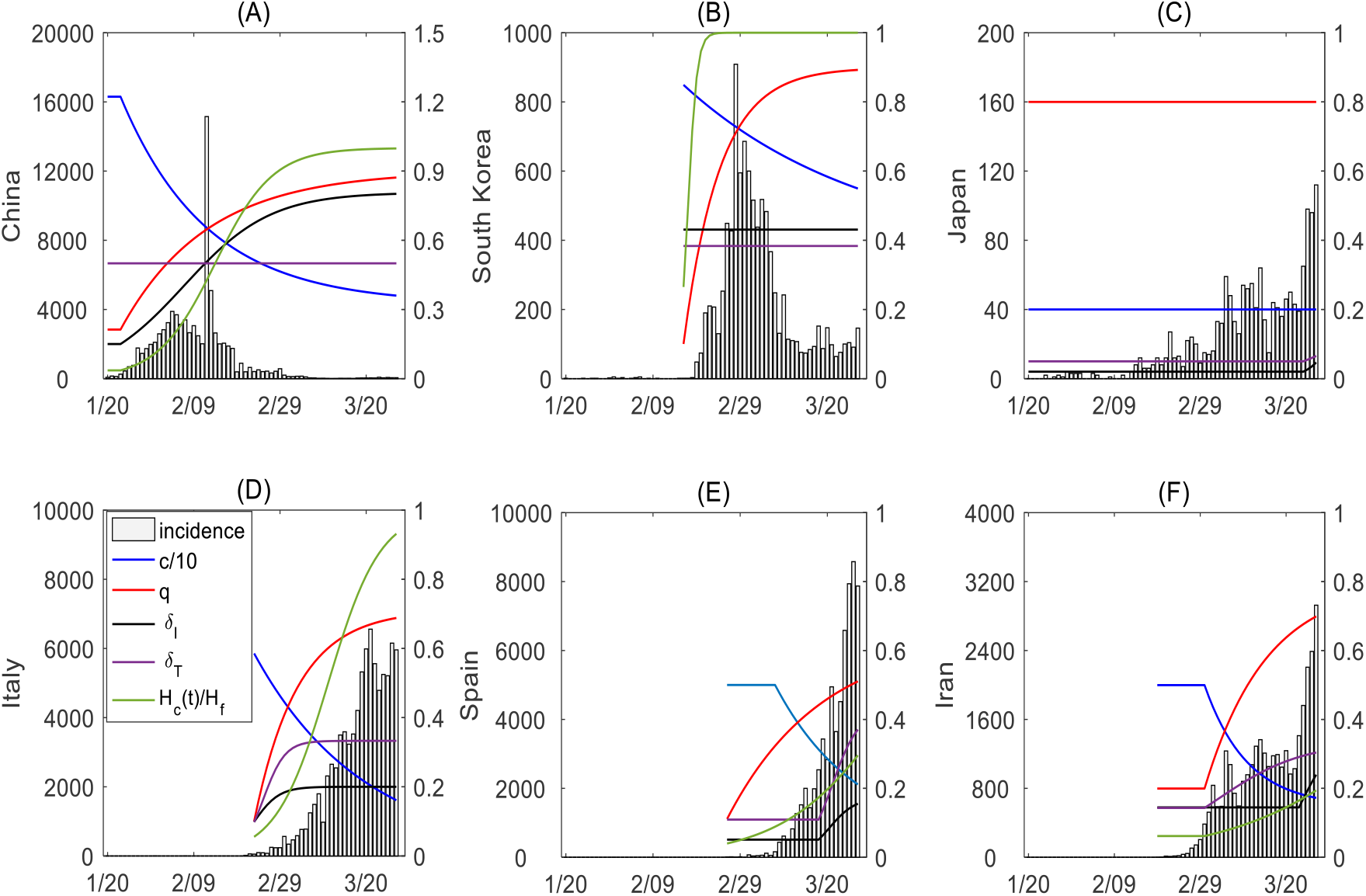
The synergistic effects of comprehensive intervention strategies and capacities of medical resources. Numbers of newly reported cases for China (A), South Korea (B), Japan (C), Italy (D), Spain (E) and Iran (F) from 23 Jan to 27 March 2020, and improving containment and mitigation measures including functions for contact rate (blue lines), quarantine rate (red), detection rate *δ*_*I*_ (black), detection rate *δ*_*T*_ (purple) and medical resources *H*_*c*_ (number of beds for each country, green). The contact rates have been divided by 10, and the numbers of beds have been divided by the carrying capacity *H*_*f*_ in each subplot.

Define *H*_*r*_ *(t)* = max{*H*_*c*_*(t)* − *H*_2_*(t)* −*θH*_1_*(t)*, 0} as the daily potential number of beds available for the individuals currently quarantined at home. Then *H*_*r*_(t) = 0 for certain days means the occurrence of a run on medical resources in some countries. In order to identify the timing of such runs for various countries, reveal the variation in the cumulative number of deaths with the detection rate, maximum number of beds and production capacity of beds, we numerically calculated *H*_*r*_(t) and the cumulative number of deaths (Fig.3 and Table 1).

**Fig.3:**
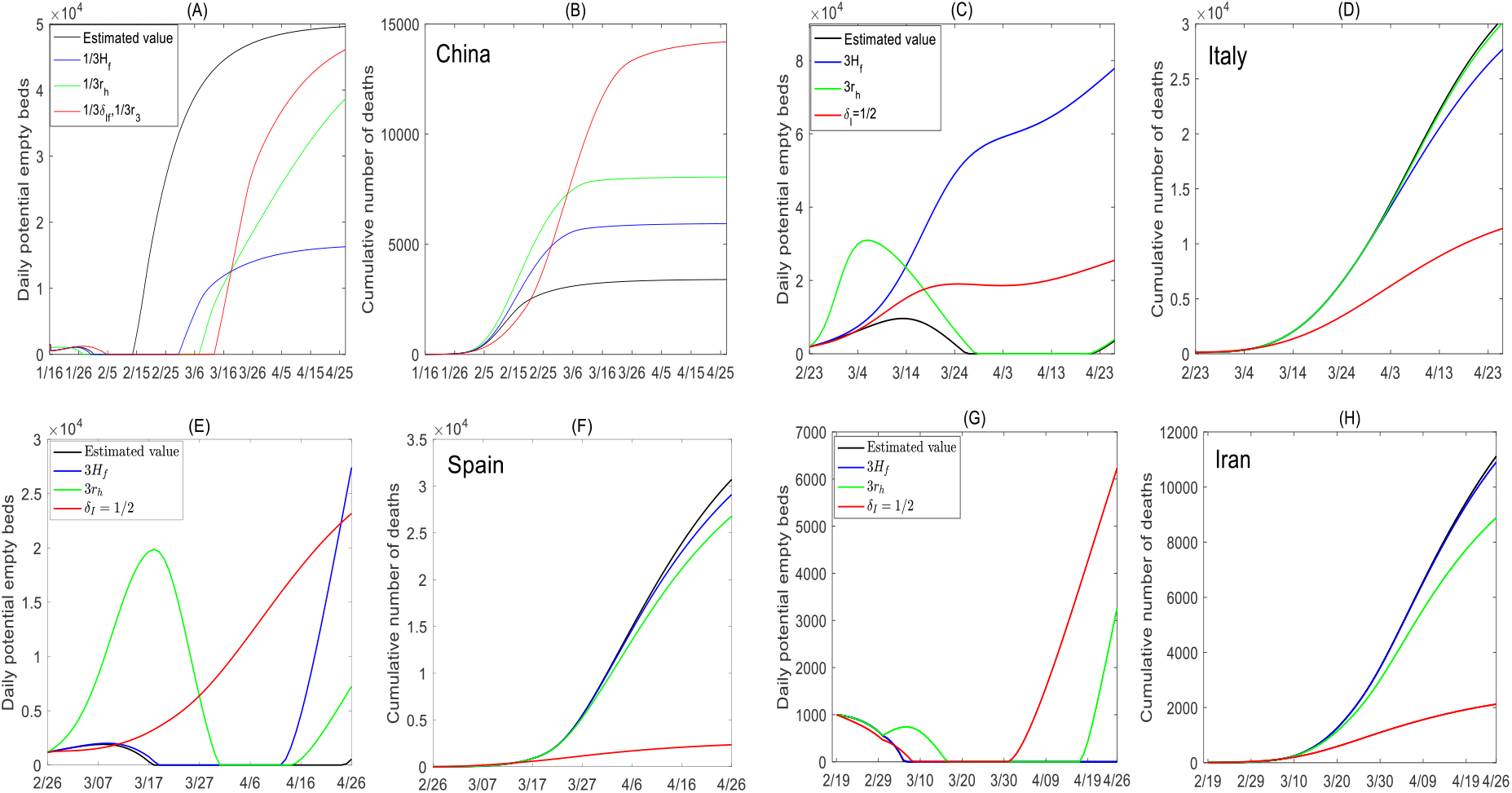
Sensitivity analyses reveal the relation between daily numbers of potential empty beds and cumulative numbers of deaths. The effects of the production capacity and reserve capacities of medical resources, and detection rates on runs on medical resources and cumulative numbers of deaths for China (A & B), Italy (C & D), Spain (E & F) and Iran (G & H). The daily potential numbers of empty beds were calculated from the formula max{*H*_*c*_*(t)* −*H*_2_*(t)* −*θH*_1_*(t)*, 0}. The black curves in each subplot are generated by the baseline parameter values listed in Table S2. The values (Table 1) of daily potential numbers of empty beds and cumulative numbers of deaths on 26 April have been calculated to reveal when and how long the runs on medical resources could last, depending on the production capacities of medical resources and detection rates for each country, when each parameter value is reduced by 1/3.

**Table 1:**
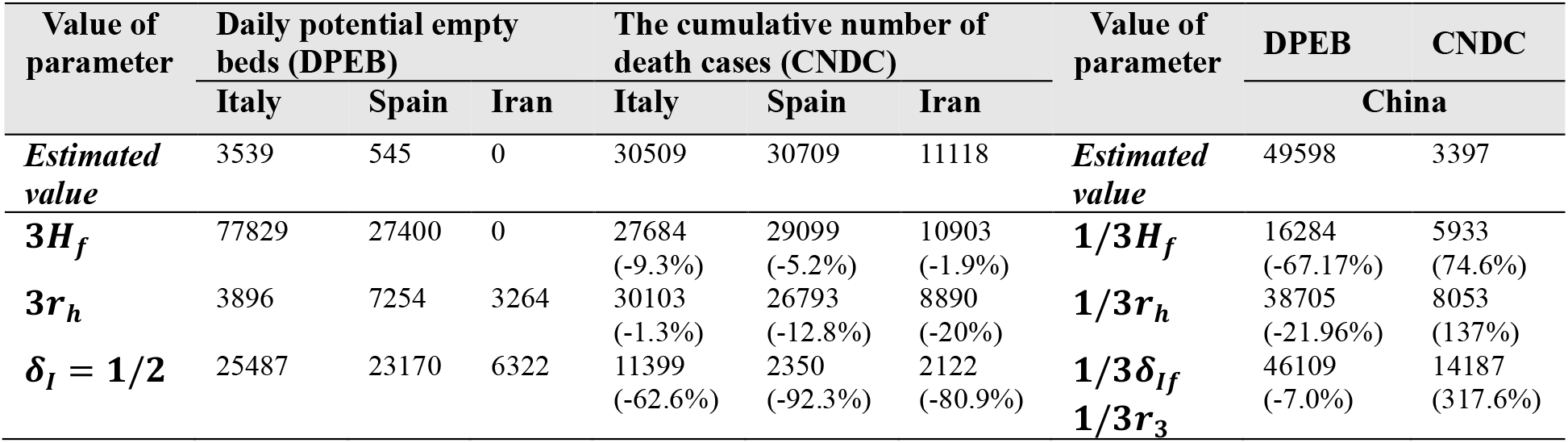
The effects of the production capacities and reserves of medical resources, and detection rates on runs on medical resources and cumulative numbers of deaths **o**n 26 April 2020.

| Value of parameter | Daily potential empty beds (DPEB) |  |  | The cumulative number of death cases (CNDC) |  |  | Value of parameter | DPEB | CNDC |
| --- | --- | --- | --- | --- | --- | --- | --- | --- | --- |
|  | Italy | Spain | Iran | Italy | Spain | Iran |  | China |  |
| *Estimated value* | 3539 | 545 | 0 | 30509 | 30709 | 11118 | *Estimated value* | 49598 | 3397 |
| $3H_f$ | 77829 | 27400 | 0 | 27684<br>(-9.3%) | 29099<br>(-5.2%) | 10903<br>(-1.9%) | $1/3H_f$ | 16284<br>(-67.17%) | 5933<br>(74.6%) |
| $3r_h$ | 3896 | 7254 | 3264 | 30103<br>(-1.3%) | 26793<br>(-12.8%) | 8890<br>(-20%) | $1/3r_h$ | 38705<br>(-21.96%) | 8053<br>(137%) |
| $\delta_I = 1/2$ | 25487 | 23170 | 6322 | 11399<br>(-62.6%) | 2350<br>(-92.3%) | 2122<br>(-80.9%) | $1/3\delta_{If}$<br>$1/3r_3$ | 46109<br>(-7.0%) | 14187<br>(317.6%) |

By calculating *H*_*r*_*(t)* with the estimated parameters, we see that the short-term shortage of medical resources in February occurred in mainland China from 1 to 13 February. However, after the specially built Huoshenshan Hospital began to treat patients on 4 February and *Fangcang* (shelter/observation ward) hospitals began to treat patients on 5 February, the number of beds increased rapidly, effectively alleviating the shortage of medical resources. However, the cumulative number of deaths would increase by 74.6% (or 137%), if the capacity (or growth rate) of beds were reduced from *H*_*f*_ to 1/3*H*_*f*_ (or *r*_3_ to 1/3*r*_3_) (Fig.3 (A-B) and Table 1). If the two parameters δ_I*f*_ (related to the detection ability) And *r*_3_ were reduced at the same time to 1/3 of the estimated values, then the duration of the run on medical resources would increase significantly (from 12 days to 37 days) and consequently the cumulative number of deaths would increase rapidly, up to 317.6% by 26 April (Fig.3 (A-B)). If the detection rate is low, the run on resources will occur later, but eventually it will occur in a wider time range, which will lead to more people becoming infected and more deaths. This indicates that rapidly increased supplies of medical resources and disease detection effectively reduced the mortality in China.

Increasing the provision of beds (3*r*_*h*_ here) threefold in Italy, Spain and Iran will only alleviate the shortage of resources for a short time, and then the run on medical resources will happen again soon afterwards (green curves in Fig.3 (C,E,G)). Italy and Spain began to recover slowly, after almost 25 days and 15 days, respectively, with zero beds remaining. Iran began to recover at a faster speed after a long period with zero beds remaining. Nevertheless, Iran had the largest reduction in the number of cumulative deaths (a reduction of 20%), followed by Spain (12.8% reduction) (Table If the maximum number of beds is only increased to 3*H*_*f*_, the results shown in Fig.3 clarify that there is little impact on the death toll in Spain and Iran (Fig.3(F, H)). Increases in the detection rate in Italy, Spain and Iran greatly reduces the cumulative number of deaths (red curves in Fig.3(D, F, H). This is because increasing the detection rate leads to significant declines in the numbers of new infections, due to strict quarantine and isolation strategies, which not only decreases the number of deaths but also avoids runs on medical resources (red curves in Fig.3(C, E)). Furthermore, in Iran increasing the detection rate still induces a run on resources in the early stage of the epidemic but leads to recovery at a later stage due to reduced numbers of new infections (black curves in Fig.3(E)), implicating the weak medical resources in Iran.

Having taken continuous and strengthened prevention and control measures, China and South Korea have quickly controlled the epidemic. The key processes for quick mitigation of the epidemic are the intensity of prevention and control measures represented by the four rates *r*_1_, *r*_2_, *r*_3_ and *r*_*h*_: the contact rate declines, quarantine rate increases, decreasing periods for detection and increasing rates of medical resource production, respectively (see SM for explanations). Reducing these rates by 10%, 20%, 30%, 40% and 50% simultaneously would have resulted in increased fractions in the cumulative numbers of confirmed cases and deaths (Table 2). Comparing the changing rates indicates that China’s control efficacy is better than South Korea’s, especially in terms of reducing deaths. Given a reduction by 50% in the comprehensive control measures in China, the cumulative number of confirmed cases and deaths would have exceeded 590000 and 60000, respectively, by 27 March. Therefore, without the very strong and comprehensive prevention and control measures that were invoked *(7, 8)*, the development of China’s epidemic would have been unimaginable, with exceedingly large numbers of cases and a surprisingly high death toll.

**Table 2:** The effects of decreasing *r*_1_, *r*_2_, *r*_3_ and *r*_*h*_ on the cumulative numbers of confirmed cases and cumulative numbers of deaths on 27 March 2020 in China and South Korea.

| Cases | The cumulative number of confirmed cases |  | The cumulative number of deaths |  |
| --- | --- | --- | --- | --- |
|  | China | South Korea | China | South Korea |
| Case $C_1$ | $8.256 \times 10^4$ | 9477 | 3329 | 170 |
| Case $C_2$ | $1.0426 \times 10^5$ (+26.3%) | 10693 (+12.8%) | 5175 (+55.4%) | 190 (+11.8%) |
| Case $C_3$ | $1.3881 \times 10^5$ (+68.1%) | 12401 (+30.9%) | 8618 (+158.8%) | 217 (27.6%) |
| Case $C_4$ | $1.9892 \times 10^5$ (+141.0%) | 14941 (+57.7%) | 15396 (+362.4%) | 251 (+47.6%) |
| Case $C_5$ | $3.1711 \times 10^5$ (+284.1%) | 18962 (+100.0%) | 30027 (+801.9%) | 297 (+74.7%) |
| Case $C_6$ | $5.9333 \times 10^5$ (+618.7%) | 25864 (+172.9%) | 63657 (+1811.9%) | 370 (+117.6%) |

Note that $r_1$ , $r_2$ , $r_3$ , and $r_h$ are the estimated values for corresponding countries, and for South Korea $r_3 = 0$ due to its constant diagnosis rate in Case $C_1$ . For Cases $C_2$ , $C_3$ , $C_4$ , $C_5$ and $C_6$ , the four rates $r_1$ , $r_2$ , $r_3$ , and $r_h$ were simultaneously reduced by 10%, 20%, 30%, 40% and 50% respectively.

It is known that the marked differences in the epidemics in various countries are associated with differences in the implementation of multiple strategies for public interventions together with increasing medical resource capacities. By estimating time-dependent contact, quarantine and detection rates we quantified enhancements to prevention and control strategies, and modeled the dynamics of medical resources, which were nested within the epidemic model proposed, revealing the timings of runs on medical resources in six different countries. Thus, the model could be used to assist health authorities to predict when they will be most in need of hospital beds, equipment such as ventilators, personal protection equipment (PPE), drugs and medical staff. The model also describes the effect of improving control measures on the complex patterns of epidemics in different countries, shows that detection rates are crucial for reducing morbidity and mortality and that synergies between prevention and control measures and medical resource availability are essential for successful control of COVID-19.

Detection of infections is a key process that significantly affects the numbers of confirmed cases and deaths. This is especially so for an increasing detection rate, which, while increasing the number of confirmed cases in the short term, leads to declines in the number of new infections. This is due to strict contact tracing followed by quarantine and isolation, and consequently reductions in the number of confirmed cases and deaths in the long term. Moreover, increasing the detection rate may result in runs on medical resources, depending on their initial capacities. As illustrated in Fig.3 (C, E and G) merely increasing detection rates in Italy or Spain did not induce runs on medical resources but these did occur in Iran (red curves). Hence the synergistic effect of improving medical capacity and production with enhancing detection is essential to mitigating the COVID-19 pandemic as well as avoiding runs on medical resources.

We suggest that Japan should pay more attention to increasing medical resources as its detection rate has increased since 25 March, otherwise the numbers of confirmed cases and deaths will increase quickly as the intensity of its control measures is not as high as in South Korea or even in Iran. If Iran had medical conditions equivalent to those of South Korea, the effects of its control measures would be far more effective and the current situation would be less severe. Comparing the estimated rate functions for Italy, Spain and Iran indicates that a low detection rate is a key process that significantly affects the epidemic, while Iran is mainly affected by its limited medical resources. Regardless of other factors, improving the detection rate can effectively and rapidly reduce the mortality rate, even after runs on medical resources. Therefore, in order to effectively reduce the numbers of new infections and mortality in COVID-19 outbreaks, detection rates should be increased while improving the production and capacity of medical resources. The synergistic effects of comprehensive prevention and control strategies are likely to succeed in mitigating epidemics, as shown by the experience of China and South Korea *(1-3,5-8)*, from whose examples other countries can learn.

## Data Availability

The data in this paper are all public data which can be found from the websites of WHO and Centers for Disease Control and Prevention of those countries.

## Supplementary Materials

SM1. Model formulation

SM2. The definition of rate functions

SM3. Comparison of the intensity of control measures based on parameter values

SM4. The data and uncertainty analysis

SM5. Table for parameter definitions

SM6. Tables for parameter estimations

## Acknowledgements

This work was partially supported by the National Natural Science Foundation of China (NSFCs: 11631012, 61772017), and by the Fundamental Research Funds for the Central Universities (GK202007001, GK202003005, xzy032020028).

## Supplementary material (SM)

This supplementary material (SM) provides detailed model description, definition of all rate functions, and comparisons of the intensities of control measures based on identified parameter values. In addition, medical resource limitations for each country are described and discussed in terms of numbers of hospital beds.

### SM1: The transmission model incorporating interventions

In order to reveal the marked differences between the applications of various control strategies independently or simultaneously under medical resource limitation, we propose a generalized COVID-19 epidemic transmission dynamics model as shown in Fig.S1 *(1,2)*:

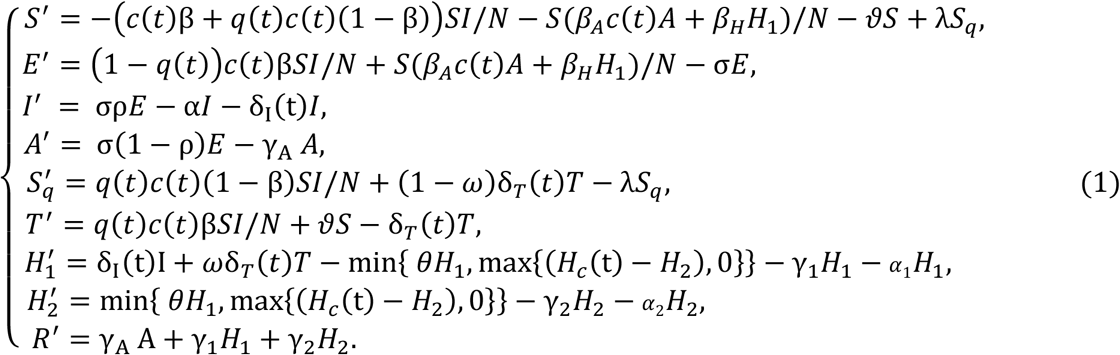

In this model, *S, S*_*q*_, *E, T, I, A, H*_1_, *H*_2_, *R* represent the populations of susceptible *(S)*, quarantined susceptible with contact tracing *(S*_q_), exposed *(E)*, quarantined suspected with contact tracing *(T)* including patients visiting fever clinics, infected *(I)* with symptoms, infected (A) but asymptomatic, people after a first medical visit but without a confirmed COVID-19 diagnosis and who are requested to quarantine at home (H_1_), hospitalized and confirmed (H_2_) and recovered (R). *N* denotes the total population, and H_c_(t) represents the capacity of hospital beds at time t which is used to describe the medical resource capacity. The detailed definitions can be found in section SM2.

In model (1), through contact tracing a proportion, q, of individuals exposed to the virus is quarantined, and can either move to compartment *T* or *S*_*q*_, depending on whether they are infected or not. The other proportion, *1 – q*, consists of individuals exposed to the virus who are missed from contact tracing and move to the exposed compartment *E* when they become infectious, or else they stay in compartment *S*. Further, we denote the transmission probability by β and the contact rate by c. Then, the quarantined individuals, if infected (or uninfected), move to compartment *T* (or S_q_) at a rate of *βcq* (or (1 – *β)cq)*). Those who are not quarantined, if infected, will move to compartment *E* at a rate of βc(1 −q). We denote by constant *ϑ* the transition rate from the susceptible class to the suspected compartment via general clinical medication due to fever or other symptoms. Meanwhile, the transmissibility of asymptomatic patients is lower than that of symptomatic patients, thus *β*_*A*_ < *β*. Further, those confirmed but not hospitalized can transmit virus to their family members, or others during their visits to health care facilities, and grocery stores etc. so *β*_*H*_ < *β*.

We note that the data on suspected individuals and also most of the confirmed cases come from the compartment *T* in China and South Korea. The suspected individuals leave this compartment at a rate δ_*T*_*(t)*, with a proportion, *ω*, confirmed to be infected by COVID-19 going to the compartment *H*_1_, whilst the other proportion, 1-*ω*, proven to be uninfected moves to the quarantined susceptible class *S*_*q*_ once recovered. People confirmed with COVID-19 may not be hospitalized due to a limitation of hospital beds, thus the term min {*θH*_1_, max {*(H*_*c*_(t) −*H*_2_), 0}} is used to describe the maximum number of newly hospitalized per day to maximize the hospital utilization, which is a piecewise function according to the relationship between *θH*_1_ and max {*(H*_*c*_(t) −*H*_2_), 0}, where *θH*_1_is the number of confirmed cases who are waiting for beds on day *t, H*_*c*_(t) is the number of hospital beds at time *t*. Therefore, max {*(H*_*c*_(t) −*H*_2_), 0} is the maximum number of remaining beds on the day. The detailed definitions of all parameters are given in Table S1.

### SM2: Definition of rate functions

As governments in various countries have been gradually strengthening their protection measures and medical resources, and in particular, providing more and more beds in response to the epidemic, we model the evolving number of beds by the logistic growth model

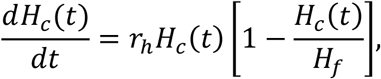

where *r*_*h*_ indicates the country-specific production capacity of medical resources in response to emerging infectious diseases and *H*_*f*_ denotes the country-specific maximum number of beds that can be provided during the disease outbreak. Therefore, these two parameters reflect the capacity of medical resources of each country in response to COVID-19 outbreaks. In the early stage of an outbreak, due to sufficient medical resources or insufficient understanding for the hospital bed needs, the number of beds is basically constant. Therefore, solving the above logistic equation, we have the number of beds on each day according to the following piecewise function:

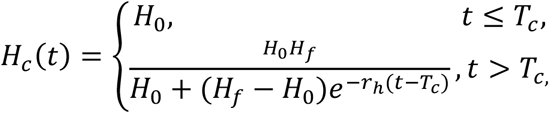

where *H*_0_ indicates the number of initial beds that can be provided to patients with COVID-19 at the beginning of the outbreak, and T_c_ denotes the critical time when each country starts to increase medical resources including hospital beds. Such critical times were 23 January and 1 March 2020 for China and Iran, respectively, while *T*_*c*_ was zero for Italy, Spain and South Korea, i.e. at the beginning of the epidemic the medical resources of these countries were constantly being supplemented. In particular, before 25 March, due to the small number of confirmed cases, Japan was not limited by any medical resources, so it is assumed that all confirmed cases were hospitalized in time or isolated at home.

Since different containment and mitigation strategies have been implemented in different countries with different critical times and intensities of strengthening prevention and control measures *(3-9)*, the functions related to contact, detection and quarantine rates in the model should be defined as piecewise functions, and the corresponding parameter values can be used to reflect the intensity at which control measures are implemented in each country. We define *c*(t), *q(t), δ*_*I*_*(t)* and *δ*_*T*_*(t)* as follows. Note that the two functions *c*(t) and *q(t)* are fixed as constants for Japan during the outbreaks, and the critical time for all rate functions defined in the following is 23 January for China, at the beginning of the epidemic for Italy.

In order to accurately describe the gradual strengthening of control strategies in this model, we assume that with increasing intensity of a control strategy the contact rate c(t) is a decreasing function with respect to time *t*, given by (1,2)

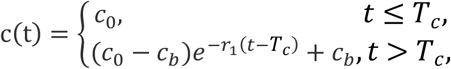

where c_0_ denotes the baseline contact rate at the initial time with c(0) = c_0_, c_b_ denotes the minimum contact rate under the current control strategies with 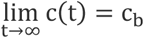, where c_b_ < c_0_, and *r*_1_ denotes how an exponential decrease in the contact rate is achieved. The critical time for Spain is 7 March *(5)*, and the critical times for other countries are the same as those in function *H*_*c*_*(t)*.

Similarly, to characterize enhanced contact tracing we define *q(t)* as an increasing function with respect to time *t*, written as

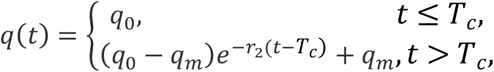

where q_0_ is the initial quarantined rate of exposed individuals with q(0) = q_0_ for China, South Korea, Italy, Spain and Iran, *q*_*m*_ is the maximum quarantined rate under the current control strategies with 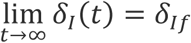 and *q*_*m*_ > q_0_, and *r*_2_ represents how an exponential increase in the quarantined rate is achieved. The critical times for each country are the same as those in function H_c_(t).

We also set the detection rate *δ*_*I*_*(t)* as an increasing function with respect to time *t*, thus the detection period 1/*δ*_*I*_*(t)* is a decreasing function of *t* with the following form:

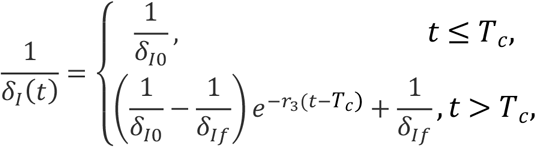

where δ_I0_ is the initial rate of confirmation, *δ*_*If*_ is the fastest confirmation rate, and *r*_3_ is the exponentially decreasing rate of the detection period. We define *δ*_*I*_(0) = *δ*_*I*0_ and 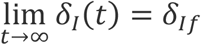 with 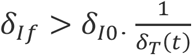 can be similarly defined. Note that both *δ*_*I*_*(t)* and *δ*_*T*_*(t)* are constants for South Korea, indicating that South Korea had a high detection rate from the beginning of the epidemic. Due to the development of the epidemic, Japan began to gradually improve its detection rate from 25 March, so the critical time for both of these detection rate functions is 25 March. The critical time for both detection rate functions *δ*_*I*_*(t)* and *δ*_*T*_*(t)* is 17 March in Spain *(5)*. Similarly, due to its limited medical resources, Iran first strengthened the detection rate of suspected cases on 1 March *(4)*, and then further expanded the detection range on 23 March, therefore the critical time for *δ*_*T*_*(t)* is 1 March, and for *δ*_*I*_*(t)* it is 23 March.

### SM3: Comparison of the intensity of control measures based on parameter values

**Detection rates** *δ*_*I*_ And *δ*_*I*_*(t), δ*_*T*_ And *δ*_*T*_*(t)*

As the definition of the number of confirmed cases from the population waiting to be tested (i.e. the *T* class) has not considered the process of passing the incubation period, the baseline parameter values of *δ*_*I*_*(t)* in some countries are greater than those of *δ*_*T*_*(t)*, as shown in Table S2. The detection rates of Japan, Italy and Spain are relatively lower than those for China and South Korea. However, Japan did not gradually improve its detection rate until 25 March, while the detection rate for Iran was increasing and finally tended to a level almost similar to that for China, which indicates that Iran is constantly improving its detection rate.

### Quarantine intensity *q* or *q(t)*

The isolation rate q in Japan has been very high since the outbreak, which fully reflects the high intensity of self-isolation in Japan, with the strong self-discipline of its citizens being one of the important factors resulting in the low level of the outbreak. The final quarantine rates of China, South Korea and Iran tend to be close to those of Japan with a relatively high growth rate R_2_. In contrast, the baseline values of quarantine rates for Italy and Spain are relative low, and the growth rate for Spain is smaller than those of all other countries.

### Contact rate *c* oR *c(t)*

There is no doubt that early in the outbreak in China, the number of contacts with susceptible persons per infected person was the largest. The final contact numbers of Italy, Spain and Iran are relatively low, which indicates that these countries have escalated social distancing measures in the later period. If they continue to maintain this level of measures for a long enough period, the outbreak can be halted. However, Japan has always adopted a relatively mild prevention and control strategy, and its exposure number has been maintained at a high level, while the limit value *c*_*b*_ of South Korea is relatively large. This reveals that although the cumulative number of reported cases in these two countries is not large at present, the number of newly reported cases may increase repeatedly in the near future. This could be particularly serious for Japan.

### Capacity of medical resources *H*_*c*_ And *H*_*c*_*(t)*

Due to the low cumulative number of confirmed cases and the low number of newly reported cases, there is no problem of limited medical resources in Japan so far, but with the development of the epidemic, whether there will be a run on medical resources or not remains to be seen. The production capacity *r*_*h*_ of medical resources in Italy is the largest except for South Korea, and then China. Compared with the cumulative number of reported cases in China and Italy, as well as the capacity to provide medical resources, i.e. the number of beds, it is clear that there is a run on medical resources or a shortage of medical resources in Italy, which is much more serious than that in China. Spain and Iran are the slowest in terms of capacity to supplement medical resources, which may also be one of the reasons for the high cumulative number of reported deaths in the two countries. It is also closely related to the recovery rate discussed below.

### Recovery rate and disease-induced death rate of isolated cases at hospital *γ*_2_ and *α*_2_

As for the recovery and mortality rates, since the epidemic data only report the number of confirmed or in-hospital cases, we can only compare the recovery and disease-induced death rates of isolated in-hospital patients here. The hospital treatment time in South Korea, Spain, Italy and China is relatively short. The mortality rate of patients in hospital is the highest in Spain, Iran and Italy, and the lowest in China. The reason for the high mortality rate in Japan may be that only severely ill patients are admitted to hospitals there.

### SM4: The data and uncertainty analyses

We obtained the numbers of daily confirmed cases, cumulative numbers of deaths and other data on COVID-19 in mainland China from NHCC *(6)*; and those in South Korea, Italy, Japan, Spain and Iran from the KCDC *(7)*, MSPC *(8)* and WHO *(9)*, respectively. The critical times for strengthening or changing national prevention and control measures can be found in publications *(3-9)*.

In order to analyze the influence of the data randomness on parameter estimation and model prediction, we assume that the epidemic data of each country follows a Poisson distribution, and we randomly generate 1000 columns of datasets for fitting. We then obtain the 95% confidence intervals for the curves generated by the real data estimations in Fig.1 and Fig.S2.

---

**Fig.S1:**
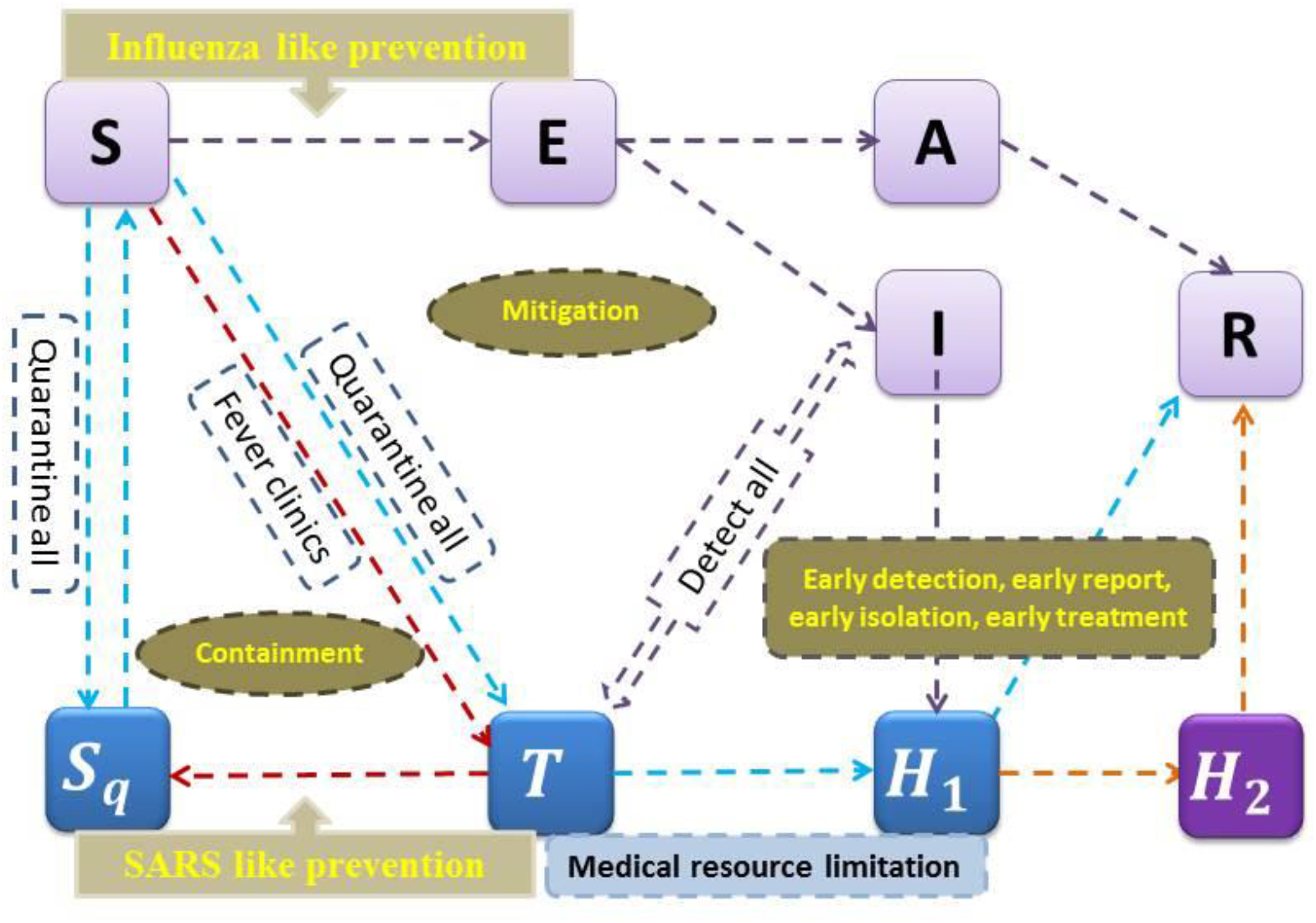
Flow diagram for the COVID-19 epidemic model incorporating containment and mitigation measures, where the medical resources limitation is described in terms of numbers of hospital beds.

**Fig.S2:**
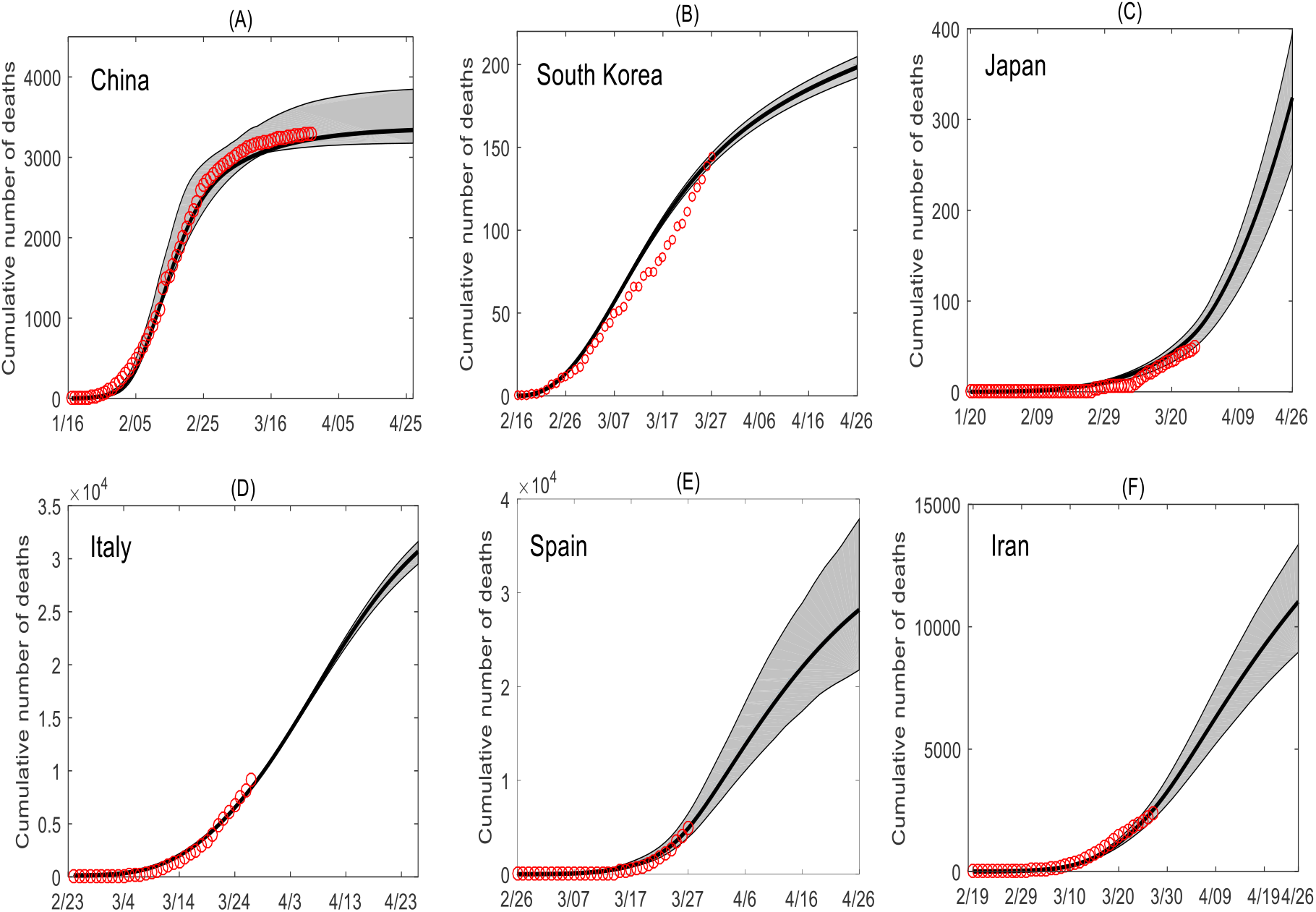
Data fitting and 95% confidence intervals of cumulative numbers of deaths for six countries indicated in each subplot.

## SM5. Table for parameter definitions

**Table S1:** Parameter definitions for the COVID-19 epidemics.

| Parameter | Definitions |
| --- | --- |
| $c(t)$ | $c_0$ Contact rate at the initial time<br>$c_b$ Minimum contact rate under the current control strategies<br>$r_1$ Exponential decreasing rate of contact rate |
| $c$ | Constant contact rate |
| $\beta$ | Probability of infected individuals transmission per contact |
| $q(t)$ | $q_0$ Quarantined rate of exposed individuals at the initial time<br>$q_m$ Maximum quarantined rate of exposed individuals under the current control strategies<br>$r_2$ Exponential increasing rate of quarantined rate of exposed individuals |
| $q$ | Constant quarantined rate of exposed individuals |
| $\vartheta$ | Quarantined rate of susceptible individuals to the suspected class |
| $\beta_A$ | Probability of asymptomatic individuals transmission per contact |
| $\beta_H$ | Probability of quarantined infected individuals transmission per contact |
| $\rho$ | Proportion of symptomatic infection |
| $\sigma$ | Transition rate of exposed individuals to the infected class |
| $\lambda$ | Rate at which the quarantined uninfected contacts were released into the wider community |
| $\delta_I(t)$ | $\delta_{I0}$ Initial diagnosis rate of infected individuals<br>$\delta_{If}$ Fastest diagnosis rate of infected individuals<br>$r_3$ Exponential increasing rate of diagnosis rate of infected individuals |
| $\delta_I$ | Constant diagnosis rate of infected individuals |
| $\delta_T(t)$ | $\delta_{T0}$ Initial diagnosis rate of quarantined individuals<br>$\delta_{Tf}$ Fastest diagnosis rate of quarantined individuals<br>$r_4$ Exponential increasing rate of diagnosis rate of quarantined individuals |
| $\delta_T$ | Constant diagnosis rate of quarantined individuals |
| $\omega$ | Confirmation ratio of quarantined exposed individuals |
| $\theta$ | Rate at which the confirmed infected individuals were hospitalized |
| $\gamma_A$ | Recovery rate of asymptomatic infected individuals |
| $\gamma_1$ | Recovery rate of confirmed infected individuals without hospitalization |
| $\gamma_2$ | Recovery rate of hospitalized infected individuals |
| $\alpha$ | Disease-induced death rate of infected individuals |
| $\alpha_1$ | Disease-induced death rate of confirmed infected individuals without hospitalization |
| $\alpha_2$ | Disease-induced death rate of hospitalized infected individuals |
| $H_c(t)$ | $H_0$ The number of available hospital beds at the initial time<br>$H_f$ Maximum capacity of hospital beds<br>$r_h$ Exponential increasing rate of hospital beds |

| Initial values | Definitions |
| --- | --- |
| $S(0)$ | Initial susceptible population |
| $E(0)$ | Initial exposed population |
| $I(0)$ | Initial infected population |
| $A(0)$ | Initial asymptomatic population |
| $T(0)$ | Initial suspected population |
| $S_q(0)$ | Initial quarantined susceptible population |
| $H_1(0)$ | Initial confirmed infected population without hospitalization |
| $H_2(0)$ | Initial hospitalized infected population |
| $R(0)$ | Initial recovered population |

## SM6. Tables for parameter estimations

**Table S2:**
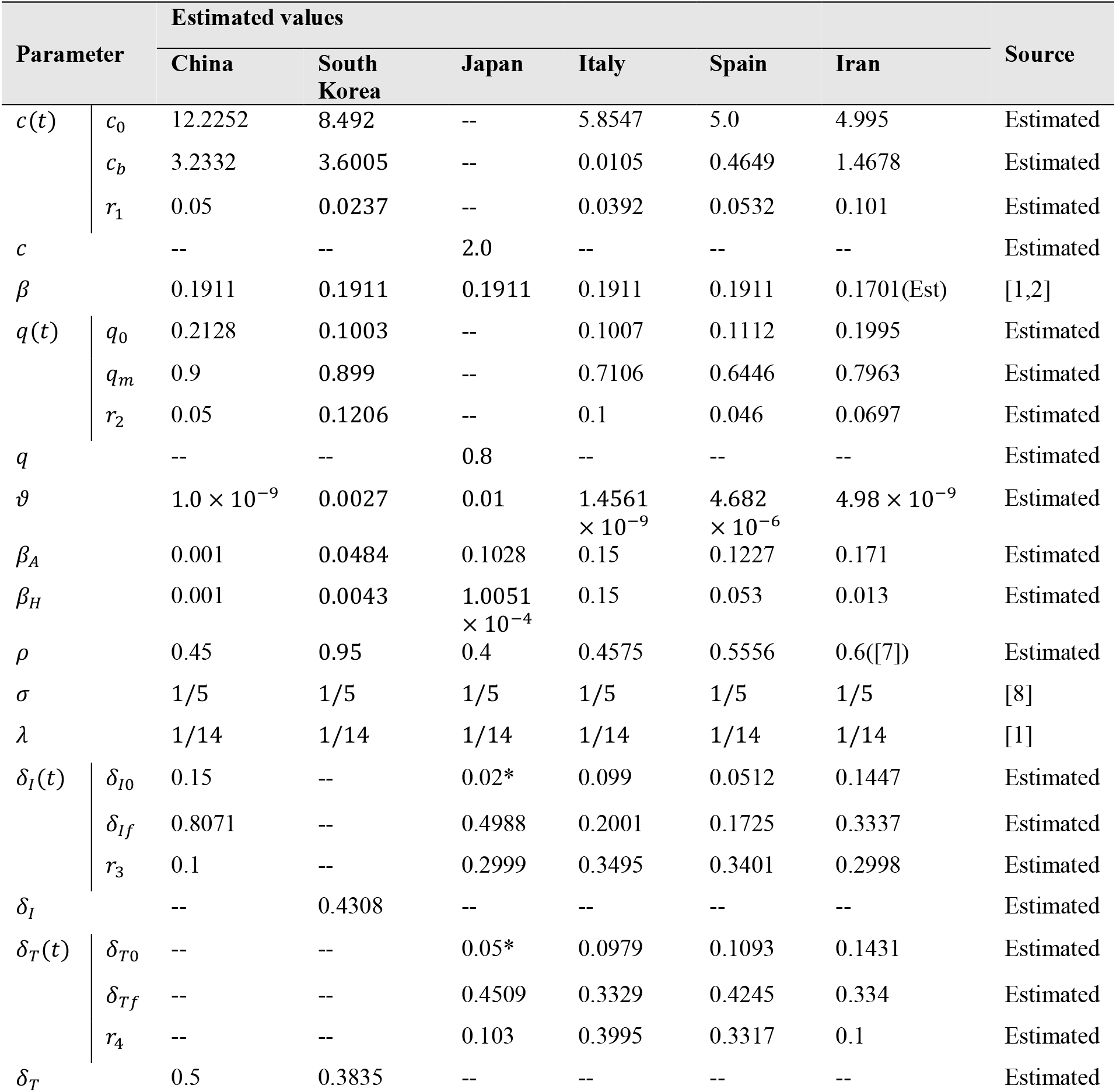
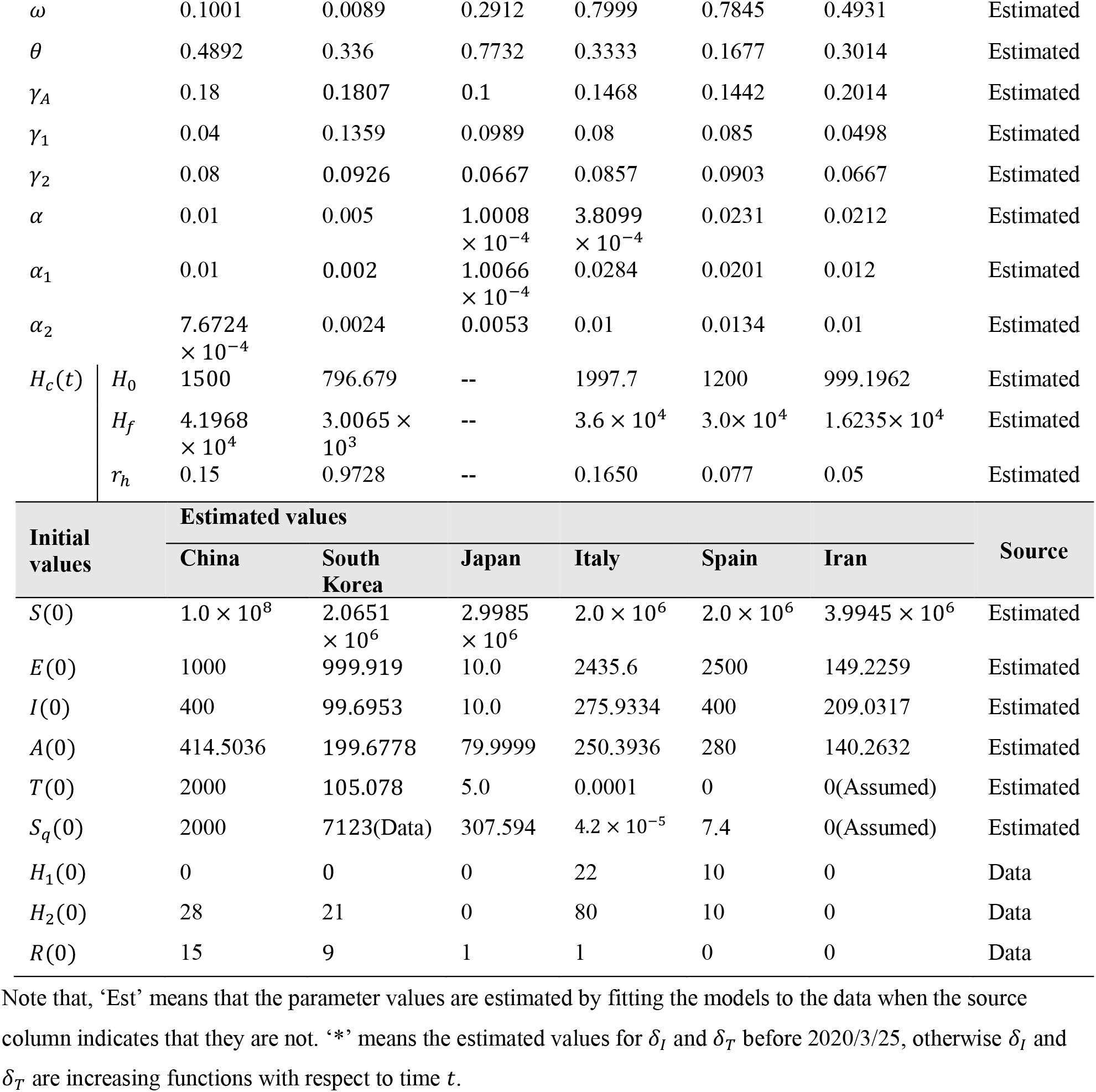
Parameter estimates for the COVID-19 epidemics in China, South Korea, Japan, Italy, Spain and Iran.

